# Explainable artificial intelligence identifies an AQP4 polymorphism-based risk score associated with brain amyloid burden

**DOI:** 10.1101/2024.02.05.24302223

**Authors:** Simone Beer, David Elmenhorst, Gerard N. Bischof, Alfredo Ramirez, Andreas Bauer, Alexander Drzezga, the Alzheimer’s Disease Neuroimaging Initiative

## Abstract

Aquaporin-4 (AQP4) is an integral component of the glymphatic system, today considered a crucial pathway for removing brain interstitial solutes like amyloid-β (Aβ). Evidence exists that genetic variation of AQP4 impacts Aβ clearance, clinical outcome in Alzheimer’s disease as well as sleep measures. We examined whether a risk score calculated from several AQP4 single-nucleotide polymorphisms (SNPs) is related to Aβ neuropathology in older cognitively unimpaired individuals. We used a machine learning approach with decision tree ensembles and explainable artificial intelligence (AI) to extract information on synergistic effects of AQP4 SNPs on brain amyloid burden from the Alzheimer’s Disease Neuroimaging Initiative (ADNI) cohort. From this information, we formulated a sex-specific AQP4 SNP-based risk score and evaluated it on the basis of data from the screening process of the Anti-Amyloid Treatment in Asymptomatic Alzheimer’s (A4) study. We found in both cohorts significant associations of the risk score with brain amyloid burden as well as amyloid positivity. The results support the hypothesis of an involvement of the glymphatic system, and particularly AQP4, in brain amyloid aggregation pathology. They suggest also that different AQP4 SNPs exert a synergistic effect on the build-up of brain amyloid burden.

## Introduction

The water channel aquaporin 4 (AQP4) is a protein which is essential for the regulation of water homeostasis in the brain (1). AQP4 is highly expressed in astrocytes, mainly polarized on their perivascular endfeet. It is considered to play a crucial role in the glymphatic system, a brain-wide perivascular network which is believed to facilitate the clearance of waste products like amyloid-β (Aβ) from the brain (2, 3). Accumulation of Aβ, followed by neuritic plaque formation, is one of the neuropathological hallmarks of Alzheimer’s disease (AD). An impaired ability to clear Aβ from the brain via the glymphatic system, the brain’s analogue to the lymphatic system of the body, could contribute to its accumulation (4). The glymphatic system is mostly active during sleep, and it was shown in mice that glymphatic influx and clearance exhibit endogenous, circadian rhythms, supported at least in part via rhythmic localization of AQP4 to the perivascular endfeet of astrocytes (5).

In a human postmortem study it was shown that AQP4 expression increased in the aging brain, and a loss of perivascular localization was significantly associated with AD status and increased Aβ burden. However, perivascular AQP4 localization was preserved among the eldest individuals who remained cognitively intact (6, 7), suggesting that a loss of perivascular AQP4 localization might contribute to the development of AD pathology. Besides glymphatic fluid clearance, increasing evidence suggests that AQP4 is also involved in brain inflammation, synaptic plasticity and memory formation, as well as regulation of extracellular space volume and potassium homeostasis (8, 9).

AQP4 is encoded by the *AQP4* gene. Genetic variation of *AQP4* may impact the expression and functionality of the water channels and therefore the clearance of Aβ. For example, Woo et al. (10) examined the functionality of one *AQP4* single-nucleotide polymorphism (SNP) in modulating the expression level of *AQP4* in an *in vitro* luciferase reporter assay, and found this SNP to cause *AQP4* expression-level changes.

Recent evidence indicates that genetic variation of *AQP4* is associated with measures of dementia. Burfeind et al. (11) have studied the effects of five *AQP4* SNPs in cognition and disease progression of AD, and have found associations with altered rates of cognitive decline after AD diagnosis. Two SNPs were associated with slower cognitive decline, two with more rapid cognitive decline, and one was associated with slower cognitive but faster functional decline. The authors also report a possible synergistic effect of the *AQP4* SNPs. Chandra et al. (12) have examined eighteen *AQP4* SNPs, and found one which was associated with decreased brain amyloid load, measured as uptake of the amyloid tracer [^18^F]Florbetapir, as well as one with increased amyloid load, disease stage progression and cognitive decline in Aβ-positive late mild cognitive impairment (MCI) and AD patients. Fang et al. (13) have studied the clinical implication of eleven *AQP4* SNPs in Parkinson’s disease (PD). They have found one SNP which was associated with slower dementia conversion, better performance in cognitive tests and lower Aβ deposition, and one SNP with faster progression to MCI and worse performance in cognitive tests. One of the SNPs also modulated an association of REM sleep behaviour disorder and the biomarker CSF Aβ42. Overall, these findings suggests that AQP4 indeed modulates cognitive disease trajectories in AD and MCI patients, potentially through moderating the build-up of disease pathology.

Recent work also describes a link between AD, sleep and AQP4. Rainey-Smith et al. (14) report an association of *AQP4* SNPs with self-reported sleep quality and a moderation of the relationship between sleep latency, duration, and brain Aβ burden. Ulv-Larsen et al. (15) showed that a haplotype of *AQP4* SNPs, containing one of the SNPs which was associated with cognition and function in (11), is also associated with slow wave energy, a combined measure of sleep intensity and duration. Despite these findings, even the latest genome-wide association studies (GWAS) did not identify *AQP4* as a risk locus for Alzheimer’s disease (16), suggesting a potential involvement of *AQP4* variants rather in endophenotypes related to AD like amyloid burden in the brain.

Furthermore, in complex diseases like AD, the disease risk is conveyed by a number of SNPs each one with small effect on disease susceptibility, which led to the formulation of polygenic risk scores (PRS) as a successful approach in AD risk prediction (17). However, the limitations of PRS are the arbitrary significance thresholds for SNP selection as well as multiple testing issues (18). In addition, PRS accounts only for the independent risk effects across multiple loci and ignores epistasis, the potentially synergistic interaction effect between genetic variants (19). However, existing strategies to explore epistasis in diseases like AD are not powerful enough due to the large number of statistical tests that need to be performed (20).

Based on these considerations, it may be hypothesized that potentially relatively small effects of single AQP4 SNPs may add up to a relevant impact in their combination. Such complex synergistic/interactive effects may be better detected by means of machine learning (ML) models as compared to conventional approaches. In contrast to the linear models used in GWAS, ML models based on decision trees like random forests or gradient boosted trees have an inherent ability to detect interactions between features, making them an excellent candidate for detecting SNP-SNP interactions even beyond only pairwise interactions (21).

Recently, a method of explainable artificial intelligence (AI) named “SHapley Additive exPlanation” (SHAP) has been introduced which computes explanations of tree-based ML models. It is based on game theory and has the ability to directly measure local feature interaction effects (22). SHAP provides also tools for understanding the global model structure based on combining the local explanations, making this method ideal for evaluating multiple SNP combinations and their importance and effects.

In this study, we tested the hypothesis that *AQP4* SNPs show a synergistic effect on brain amyloid burden. We used a parallel tree boosting method, XGBoost (“eXtreme Gradient Boosting”) (23) with the minor allele carrier status of *AQP4* SNPs as features, to model interactions between SNPs as well as their individual effects on brain amyloid burden, with data from 324 participants of the Alzheimer’s Disease Neuroimaging Initiative (ADNI) database (adni.loni.usc.edu). We used explanations derived from SHAP to formulate a SNP-based risk score, considering protective as well as deleterious effects of the SNPs, and evaluated the risk score using a causal modelling approach (24) based on multiple linear regression both in ADNI as well as in an independent cohort, data from 2987 cognitively unimpaired participants of the screening process of the A4 study (a4study.org) (25).

## Materials and Methods

Data used in this study were obtained from the Alzheimer’s Disease Neuroimaging Initiative (ADNI) database (adni.loni.usc.edu). The primary goal of ADNI was to test whether neuroimaging (serial magnetic resonance imaging (MRI), positron emission tomography (PET)) together with biological markers, clinical and neuropsychological assessment can be combined to measure the progression from cognitively normal (CN) to mild cognitive impairment (MCI) and Alzheimer’s disease (AD). In 2013, whole genome sequencing (WGS) was performed on 818 participants (26).

Further, we used data of the Anti-Amyloid Treatment in Asymptomatic Alzheimer’s (A4) study (a4study.org). The A4 study is a secondary prevention trial in preclinical Alzheimer’s disease, aiming to slow cognitive decline associated with brain amyloid accumulation in clinically normal older individuals (25). Cognitively unimpaired older individuals were selected as participants, based on evidence of amyloid accumulation on screening PET scans. Participants without evidence of elevated amyloid accumulation were eligible to screen for the Longitudinal Evaluation of Amyloid Risk and Neurodegeneration (LEARN) Study. This companion observational study serves as a longitudinal comparison group with the treatment and placebo Aβ+ arms randomized in the A4 study (27). We used the A4/LEARN pre-randomization data acquired in the screening process with imputed genetic data as from November 23, 2020.

### Study participants

From the ADNI cohort, we considered only participants with whole genome sequencing (WGS) data of *AQP4* SNPs as well as completed [^18^F]Florbetapir PET scans available. Furthermore, to ensure an ethnically homogeneous group, we considered only the 97% of the participants with ethnic category declared as “Not Hispanic or Latino”.

Depending on the period since their enrollment, ADNI participants had several follow-up visits. At the time of enrollment, the diagnosis was not exceeding mild or early forms of dementia. Therefore, we decided to use data from the last visit available, to ensure that we consider the maximum bandwidth of disease from cognitively normal to severe forms of AD. Consequently, participants were older on average and were more often diagnosed with AD compared to baseline. Overall, N= 324 participants were included.

From the A4/LEARN cohort, we also considered only participants with data of *AQP4* SNPs available, and a completed [^18^F]Florbetapir PET scan. We also restricted the ethnicity to “Not Hispanic or Latino”, which made up 99.3% of the data. Overall, N= 2987 participants were included.

For the assessment of significant differences in age of the two cohorts we performed an independent samples t-test using SciPy (https://scipy.org). For the assessment of significant differences in the percentage of APOE ε4 carriers and Aβ positives we used a chi-square test of independence of variables, also using SciPy.

### *AQP4* SNPs

Candidate *AQP4* SNPs were collected from the database of genetic variation of the National Center for Biotechnology Information (NCBI), dbSNP (28) (https://www.ncbi.nlm.nih.gov/snp/). 31 *AQP4* SNPs were selected based on their availability in both ADNI and A4/LEARN after quality control procedures which included a minor allele frequency of greater than 5%, and Hardy-Weinberg equilibrium (p < 0.001), using PLINK 2.0 (29) (https://www.cog-genomics.org/plink/2.0). We identified 3 groups of SNPs with high correlation (Pearson correlation coefficient, r > 0.96) and kept one SNP as representative for a group which either had a previously reported association in the context of AD or had the highest correlations with all other group members. Finally, 19 SNPs were used for the analysis. The decision to keep correlated SNPs up to an r^2^ of 0.92 was based on evidence that SNP-SNP pairs in high linkage disequilibrium can be significantly associated with phenotypes due to epistatic effects (30), so that cases where two correlated SNPs differ from each other might contain valuable information. Details of the SNPs which were used, as well as previously reported associations, are shown in Table 1. Figure 1 shows an illustration of the position of the SNPs within the AQP4 gene. With our SNP selection we cover the entire locus of the AQP4 gene.

**Table 1.** AQP-4 SNPs investigated in the current study.

| SNP | Consequence | Previously reported associations |
| --- | --- | --- |
| rs162005 | Intron |  |
| rs9961118 | 2KB upstream |  |
| rs72878794 | 2KB upstream | - Amyloid burden and clinical outcome in AD (12)<br>- Sudden infant death syndrome (31) |
| rs151244 | 2KB upstream | - Amyloid burden and clinical outcome in AD (12) |
| rs162006 | 2KB upstream |  |
| rs2075575 | 2KB upstream | - Parkinson’s disease (32)<br>- Leukoaraiosis (33)<br>- Sudden infant death syndrome (34, 35) |
| rs72878787 | 2KB upstream |  |
| rs162008 | 5’-UTR | - Brain plasticity and learning (10)<br>- Slow wave energy regulation in human NREM sleep (15) |
| rs3875089 | Intron | - Moderation of the relationship between sleep and brain Aβ-amyloid burden (14)<br>- Traumatic brain injury outcome (36)<br>- Cognitive decline after AD diagnosis (11) |
| rs4800773 | Intron |  |
| rs3763040 | Intron | - Cognitive decline after AD diagnosis (11)<br>- Cognitive performance in Parkinson’s disease (13) |
| rs162009 | Intron | - Cognitive performance (13) and brain activity (37) in Parkinson’s disease |
| rs35248760 | Synonymous | - SNPs with high correlation: rs72878776, rs67207056, rs60565102, rs55875625, rs12968026, rs11661256(all Intron), rs1058427 (3'-UTR)<br>- rs72878776 moderates the relationship between sleep and brain Aβ-amyloid burden (14)<br>- rs1058427 associated with intracerebral haemorrhage (ICH) and perihematomal oedema (PHE) volume (38) |
| rs71353406 | Intron | - SNPs with high correlation: rs14393, rs3763043 (all 3'-UTR)<br>- Moderation of the relationship between sleep and brain Aβ-amyloid burden (14)<br>- Traumatic brain injury outcome (36) (rs3763043)<br>- Cognitive performance in Parkinson's disease (13) (rs3763043)<br>- Cognitive decline after AD diagnosis (11)<br>- Schizophrenia in the Southern Chinese Han population (39) (rs3763043) |
| rs71353405 | Intron |  |
| rs335930 | Intron | - Slow wave energy regulation in human NREM sleep (15) |
| rs1058424 | 3'-UTR | - Schizophrenia in the Southern Chinese Han population (39) |
| rs7240333 | 3'-UTR | - Cognitive performance in Parkinson's disease (13) |
| rs16942851 | 500B<br>Downstream | - SNPs with high correlation: rs455671, rs335931 (all Intron), rs335929 (3'-UTR)<br>- Slow wave energy regulation in human NREM sleep (15)<br>- More rapid decline in Clinical Dementia Rating, but slower decline in Logical Memory and Digit Symbol tests performance (11) (rs335929) |

**Figure 1.**
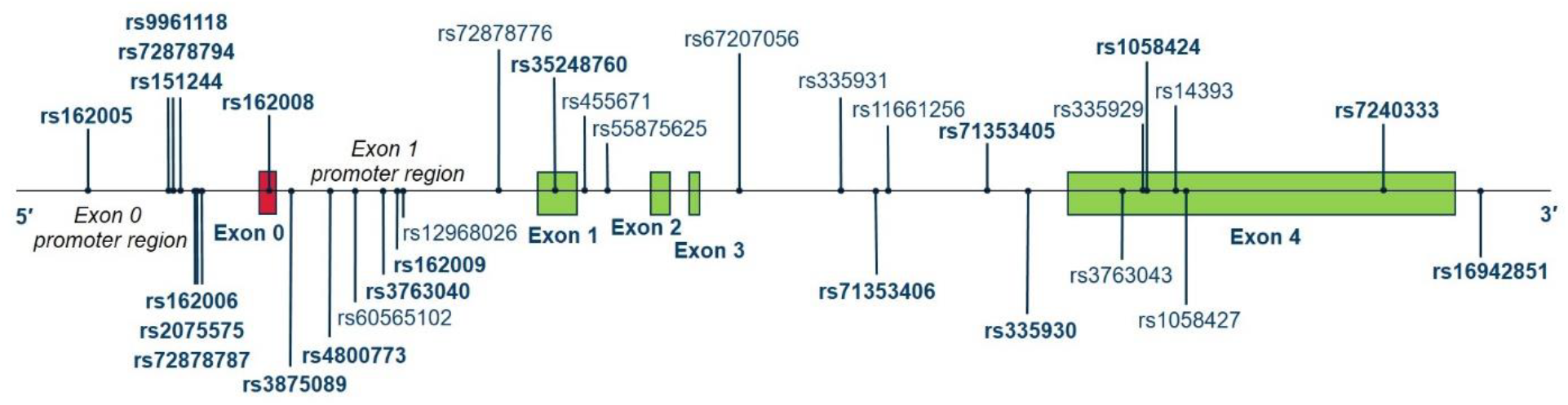
Illustration of the SNP positions within the *AQP4* gene. The 19 SNPs which were used for the analysis are represented in bold. The SNPs represented in regular type are in high correlation with one of the SNPs in bold (see Table 1).

Finally, we assigned a “carrier” status to subjects which are either homozygous in the minor allele or heterozygous, and a “non-carrier” status to subjects which are homozygous in the common allele.

### Amyloid PET imaging

For the ADNI cohort, brain amyloid burden was assessed using a region of interest based analysis of [^18^F]Florbetapir (formerly AV45) PET imaging data, which is available in a spreadsheet format from ADNI (40). In short, after the injection of approximately 10 mCi as an intravenous bolus, subjects are scanned from 50 to 70 minutes after injection in time frames of 5 minutes each. Quality control and preprocessing of the imaging data included visual inspection, correction for motion between the time frames, averaging of the motion corrected time frames to a single frame, rigid coregistration to a standard grid and intensity normalization using a reference region. To account for the various PET scanners used in the study, data were homogenized using a scanner-specific smoothing kernel. From the available spreadsheet, we used the summary [^18^F]Florbetapir cortical standardized uptake value ratios (SUVR) normalized by whole cerebellum. From these values, global Aβ positivity in individual subjects was determined by applying a cutoff value of 1.11, as described in the internal ADNI “Amyloid PET Processing Methods” document.

For the assessment of brain amyloid burden in the A4/LEARN study, participants also received an injection of 10 mCi [^18^F]Florbetapir and were scanned 50 to 70 minutes after injection. Amyloid burden is also measured as mean cortical SUVR with a whole cerebellar reference region, and is available as a spreadsheed from the A4/LEARN study. To maintain consistency with ADNI data, Aβ positivity was determined by applying a general cutoff value of 1.11, as opposed to the A4/LEARN procedure where SUVR values between 1.10 and 1.15 were considered as amyloid positive only when a visual read was also deemed positive by a consensus determination involving two readers (27).

### Feature engineering and estimation of risk score

Decision tree ensembles are ideal to capture complex data dependencies. To detect synergistic or interaction effects of the AQP4 SNPs we have used sex, APOE4 status expressed as number of ε4 alleles, and the status of the 19 AQP4 SNPs from ADNI as features, as well as brain amyloid burden as target for a regression task using XGBoost 1.7.6 (“E**x**treme **G**radient **B**oosting”) (23) (https://github.com/dmlc/xgboost). This algorithm uses many decision trees as weak learners and combines them to a strong learner by minimizing a loss function. It is known as a robust algorithm with excellent performance in a variety of applications. We have used the data from the ADNI cohort for this step because ADNI has an equal share of amyloid positive and negative participants and covers all stages of AD, from cognitively normal to dementia, in contrast to A4/LEARN with only cognitively unimpaired participants.

The prevalence of Alzheimer’s disease is higher in women compared to men (41). Moreover, AQP4 gene regulation has been found to be affected by sex-biasing factors (42), and it was also shown that AQP4 regulates the effects of ovarian hormones on neurotransmission (43). Therefore, we decided to perform the modelling separately for women and men to interrogate potential biological sex differences.

Next, we used the TreeExplainer of the SHAP package (22, 44) (https://github.com/slundberg/shap, version 0.42.0) to explain the model decisions. The TreeExplainer can explain each prediction with respect to local feature importance and feature effect towards the predicted value, as well as feature interaction effects. Local explanations can also be combined to understand the global model structure. After visual inspection of the explanations, we assigned the carrier status of an AQP4 SNP as “protective” when the global feature effect consistently leaned towards lower predicted amyloid values, and as “deleterious” when the global feature effect pointed consistently towards higher predicted amyloid values. Finally, we counted the number of deleterious SNPs for each individual to calculate a sex-specific SNP-based risk score both in ADNI and in A4/LEARN, as part of a feature engineering process where new features which are more relevant to the target can be created from existing features using domain knowledge and data analysis.

### Causal modelling and statistical analysis

Usually, machine learning modelling is performed as prediction modelling which seeks to accurately predict the outcome of interest from a set of given features, preferably with high predictive performance. However, good features for predictive modelling are often rather symptoms of the outcome than causes. For example, James et al. (45) have found that ML models outperform existing dementia risk prediction models with only six key features, namely level of independence, clinical judgement of various aspects of decline, time to complete trail making test part B, clinical dementia rating (CDR) home and hobbies impairment, CDR memory and CDR orientation. These features have a strong predictive potential for dementia, but cannot be considered as causal for the disease.

In contrast to predictive modelling, the goal of causal modelling is to estimate the true causal association between a particular variable and the outcome of interest. While randomized control trials are the gold standard to assess causal associations, they also can be studied to a certain extent in observational data by the formulation of a causal model of confounding and a careful consideration of the covariates based on this model (24, 46).

To represent our assumptions regarding the relationship between AQP4 status expressed as SNP-based risk score, brain amyloid burden measured as [^18^F]Florbetapir SUVR, and known demographic and genetic risk factors (age, sex, and APOE4 status), we developed a graphical causal model as a directed acyclic graph (DAG) (47) using DAGitty (http://dagitty.net/) (Figure 2). This causal model allowed us to assess which of the covariates have to be controlled for in a regression model. AQP4 status is the exposure of interest, brain amyloid burden the outcome. APOE4 status, age and sex affect brain amyloid burden directly, which includes all unknown or unobserved mediators. We also cannot exclude a possible effect of age and sex on both AQP4 and APOE4 expression. In (12), no relationship between AQP4 SNPs and APOE4 status could be established. Our own correlation analysis did also show no relationship, so that we did not include a relation between AQP4 and APOE ε4 in figure 2.

**Figure 2.**
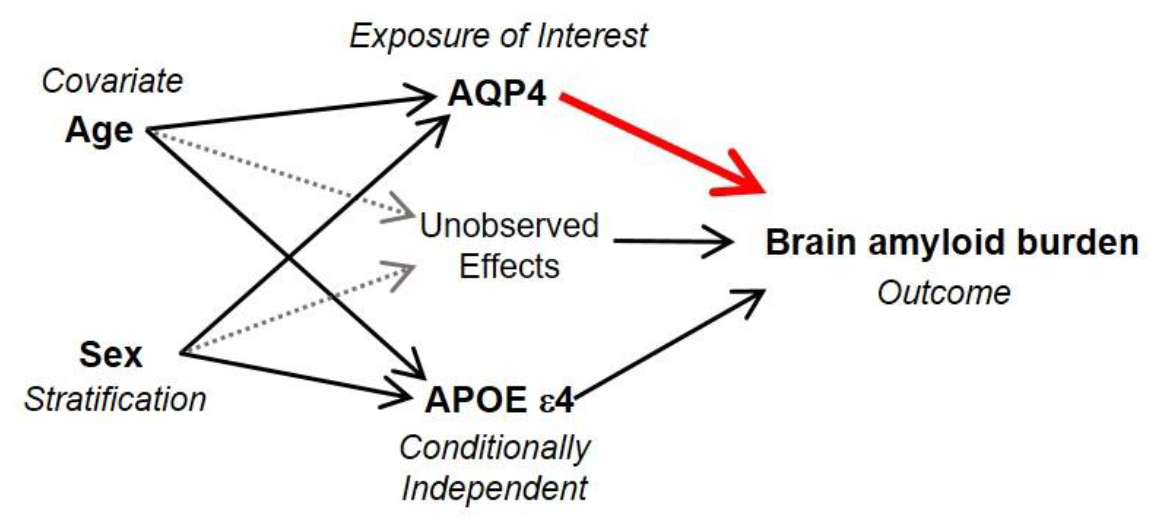
Causal model as directed acyclic graph (DAG), to assess exposure-outcome adjustment set.

According to this causal model, the minimal sufficient adjustment set for estimating the total effect of AQP4 status, expressed as SNP-based risk score, on brain amyloid is age and sex. AQP4 status is then conditionally independent from APOE4 status, so that we don’t have to control for it.

We stratified the sample on sex by analyzing the data separately within groups of males and females. Finally, we included age as a covariate in a multiple linear regression model using ordinary least squares (OLS) regression within statsmodels (https://www.statsmodels.org) for both ADNI and A4/LEARN data.

To assess an association of the risk score with the prevalence of amyloid positivity, we grouped the study participants according to their risk score and compared the groups with respect to amyloid positivity using the Cochran–Armitage trend test of statsmodels. To compare amyloid burden between groups of the same risk score, we performed an independent samples t-test using SciPy (https://scipy.org).

For all statistical tests we used p < 0.05 as the threshold for statistical significance.

## Results

### Study demographics

Table 2 shows demographic information and further characteristics for the two cohorts. The A4/LEARN cohort is significantly younger on average, and has no participants with MCI or AD. It has also significantly less APOE ε4 carriers and Aβ positives.

**Table 2.** Demographic information and clinical characteristics of study participants.

|  | ADNI | A4/LEARN | p value |
| --- | --- | --- | --- |
| Number of subjects | 324 | 2987 |  |
| Age, Mean (SD) [y] | 76.06 (6.98) | 71.36 (4.75) | **7E-56** |
| Males / Females | 183 / 141 | 1206 / 1781 |  |
| Diagnosis: Cognitively unimpaired | 125 | 2987 |  |
| Diagnosis: Mild cognitive impairment | 132 | - |  |
| Diagnosis: Dementia | 64 | - |  |
| Diagnosis: non specified | 3 | - |  |
| % APOE $\epsilon 4$ carriers | 39.5 | 35.8 | **0.0008** |
| % Aβ positive | 50.6 | 32.6 | **1E-10** |

### Feature engineering and estimation of risk score

Figure 3 shows the explanations for the global model structure as estimated by the TreeExplainer of SHAP for both women (left) and men (right) for ADNI data. The SHAP value on the x-axis represents the impact on the prediction of the model, the predicted amyloid burden. The ordering of the features from top to bottom reflects the feature importance. As could be expected, APOE4 status is explained as the most important feature. Non-carrier status of the ε4 allele (blue dots) leads to low predicted amyloid burden, and homozygous status of ε4 (red dots) leads to higher predicted amyloid burden than heterozygous status of ε4 (purple dots).

**Figure 3.**
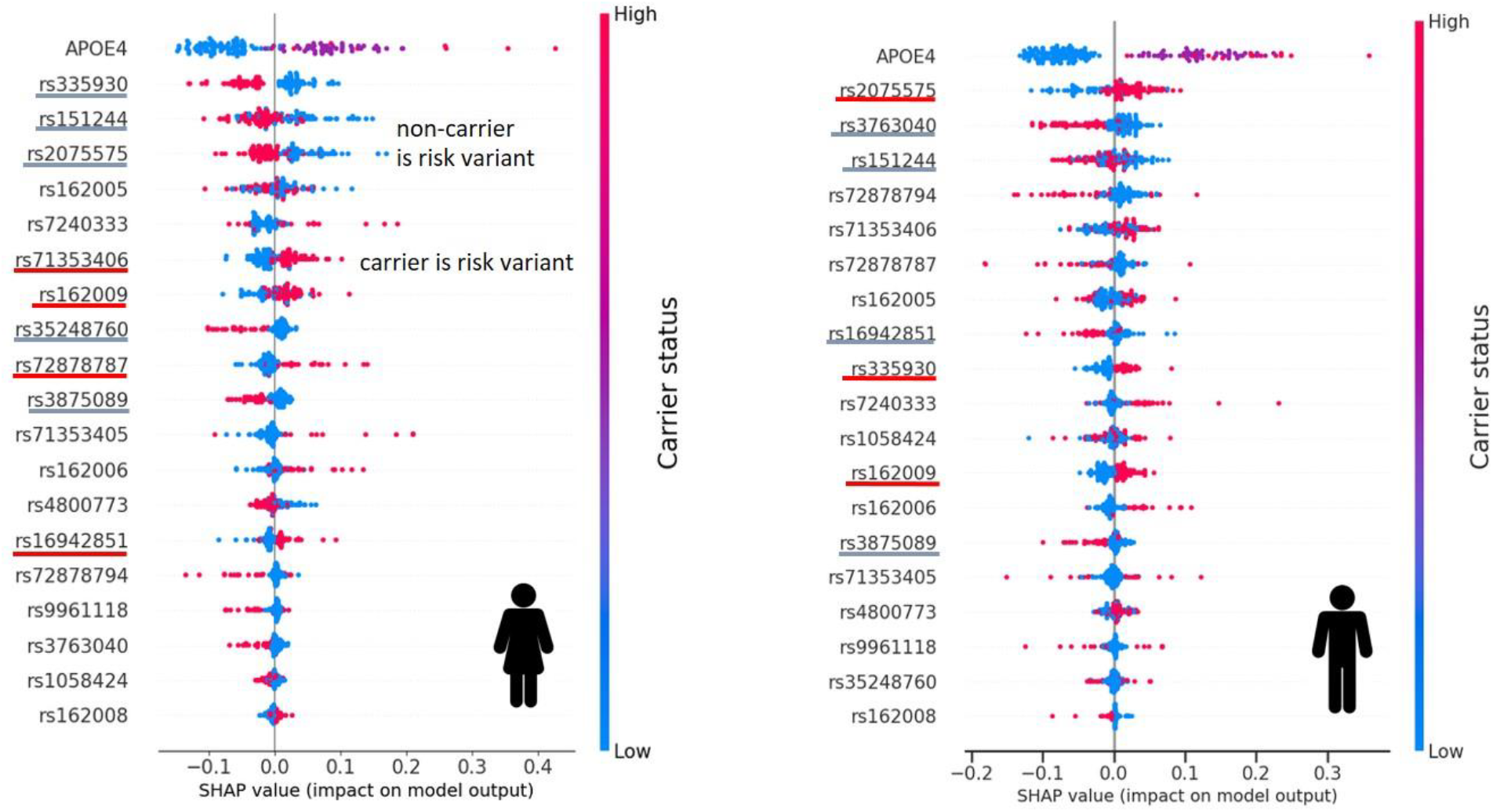
The SHAP summary plot explains the global structure of the decision tree ensemble model for women (left) and men (right). SHAP values on th x-axis represent the impact of a feature on the model’s prediction. Each dot of the beeswarm plot corresponds to an individual in the ADNI dataset. For AQP4 SNPs, red dots represent carriers (carrier status “high”) and blue dots represent non-carriers (carrier status “low”) of the SNPs. For the APOE4 feature, the colour of the dots represent the number of the *ε4* alleles (0/1/2). The SNP-based risk score is calculated from the SNPs which are underlined in blue and red, so that for the blue underlined SNPs the non-carrier is the risk variant and for the red underlined SNPs the carrier is considered as the risk variant.

The explainer also assigns for several SNPs (in total 9 for women and 7 for men) the carrier status consistently to lower or higher predicted amyloid burden. Consequently, we assigned for the blue underlined SNPs in figure 3 (rs335930, rs151244, rs2075575, rs35248760 and rs3875089 for women, rs3763040, rs151244, rs16942851 and rs3875089 for men) the non-carrier status as deleterious, for the red underlined SNPs (rs71353406, rs162009, rs72878787 and rs16942851 for women, rs2075575, rs335930 and rs162009 for men) the carrier status. There were no cases in which the selection of SNPs in an individual were all either protective or deleterious, so that counting the number of deleterious SNPs for each individual resulted in risk scores between 2 and 7 for women in the ADNI cohort and between 1 and 7 in the A4/LEARN cohort, and between 2 and 6 for men in ADNI as well as A4/LEARN. Figure 4 shows that the new feature “AQP4 SNP-based risk score” is relevant to the target for both the ADNI and the A4/LEARN cohort. For both cohorts and both males and females, the t-test shows that the differences in amyloid burden between the groups of lowest and highest risk factor are significant (ANDI females: p = 0.0018, males: p = 0.035; A4/LEARN females: p = 0.043, males: p = 0.035).

**Figure 4.**
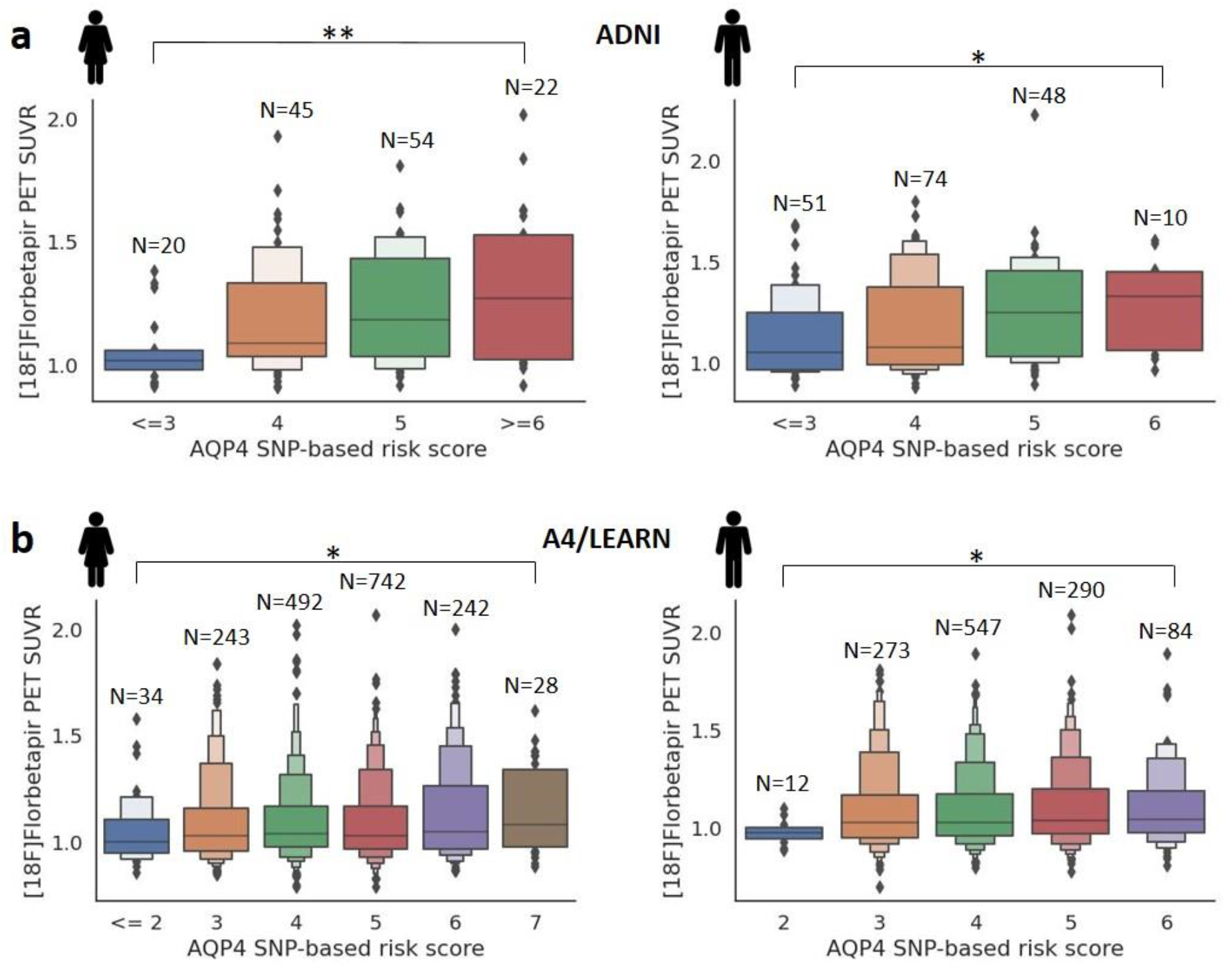
(a) In the ADNI cohort the new feature “AQP4 SNP-based risk score”, calculated as the sum of deleterious variants, is clearly relevant to the target, amyloid burden measured as ^[18F]^Florbetapir PET SUVR, for both women and men. (b) In the A4/LEARN cohort, an increase in amyloid burden can also be observed with increasing risk score. The increase is especially driven by the extreme low and high risk scores, and is more pronounced for women.

### Causal modelling and statistical analysis

#### Association between AQP4 SNP-based risk score and brain amyloid burden

For women, multiple linear regression with age as covariate showed a significant association of risk score with amyloid burden for the ADNI (p = 0.001, regression coefficient = 0.0473, 95% confidence interval: 0.02 - 0.074) as well as for the A4/LEARN (p = 0.014, regression coefficient = 0.011, 95% confidence interval: 0.002 - 0.02) cohort. For men, a significant association could be shown in ADNI data (p = 0.005, regression coefficient = 0.0552, 95% confidence interval: 0.017 - 0.094) but not in A4/LEARN data (p = 0.102, regression coefficient = 0.0106, 95% confidence interval: -0.002 - 0.023). However, for both women and men in ADNI as well as in A4/LEARN, the mean amyloid burden differs significantly between the groups with the lowest and highest risk factors (ADNI: women p=0.002, men p=0.035; A4/LEARN: women p=0.043, men p=0.035).

The distribution of PET SUVRs for the different risk scores in figure 4 suggests that the increase in average amyloid burden is not driven by a general, small increase for all subjects with this risk score, but rather by a higher number of subjects with considerably elevated amyloid burden, compared to the groups with lower risk score. This should reflect in an increase in the share of amyloid positives with increasing risk score.

#### Association between AQP4 SNP-based risk score and amyloid positivity

Fig. 5 shows that the risk score is indeed predictive for the prevalence of amyloid positivity, with a trend towards higher prevalence with increasing risk score. The Cochran-Armitage trend tests show that the trends are significant for both cohorts in women (ADNI: p = 0.002, A4/LEARN: p = 0.04) as well as in men (ADNI: 0.0007, A4/LEARN: 0.03).

**Figure 5.**
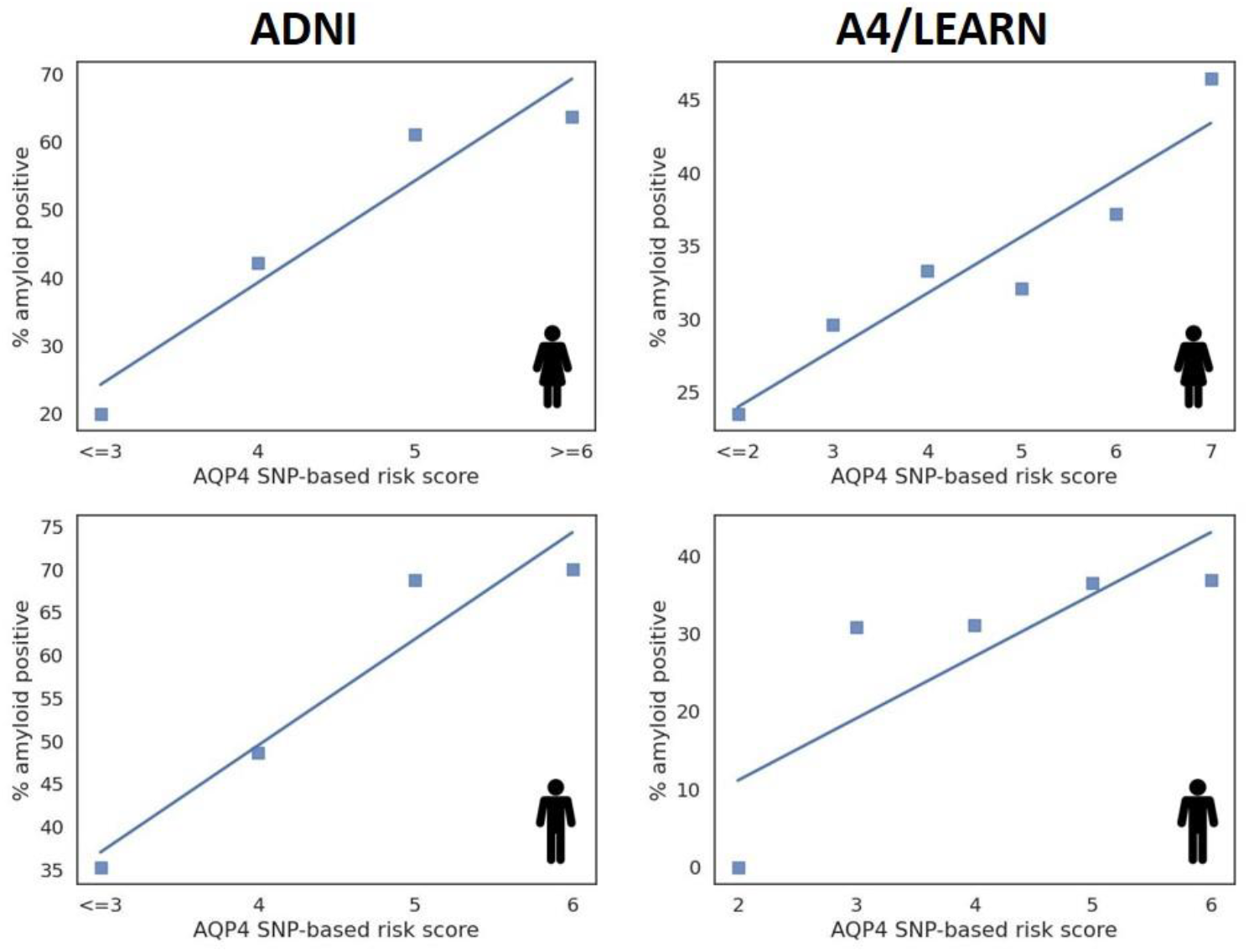
The share of amyloid positives increases with higher risk scores for both males and females in the ADNI as well as in the A4/LEARN cohort.

## Discussion

In this work, we have used decision tree ensembles and explainable AI to assess the impact of interacting *AQP4* polymorphisms on brain amyloid burden. This allowed us to define a sex-specific group of SNPs with the respective information on protective or deleterious effects, which together showed a highly significant association with brain amyloid burden and amyloid positivity. Of note, the risk score which was estimated based on ADNI data was also significant in the completely independent A4/LEARN cohort of cognitively unimpaired participants with respect to both amyloid burden and share of amyloid positivity for women, and with respect to the share of amyloid positivity also for men. For some of the individual SNPs which make up the risk score, effects on brain amyloid burden, cognition and functional decline in AD have already been described in previous work. Chandra et al. (12) found that the carrier status of rs151244 was associated with increased Aβ uptake, but only for Aβ positive patients with late MCI or AD. Burfeind et al. (11) found that the carrier status of rs3875089 was associated with slower cognitive decline or dementia progression. They also found that rs3763040 and rs3763043, which is in strong linkage disequilibrium (LD) with rs71353406, were associated with faster cognitive decline or dementia progression.

However, they found also an inconsistent association of rs335929, which is in strong LD with rs16942851, with slower cognitive but faster functional decline. Similarly, Fang et al. (13) found further inconsistent associations of rs3763040 and rs3763043 with worse executive but better visuospatial function in a cohort of PD patients. The reason for these discrepant findings might be the occurance of a statistical peculiarity named Simpson’s paradox, which describes the phenomenon that a trend observed in the overall data can disappear or even reverse when the data are stratified or analyzed with respect to subgroups. This effect is the consequence of an extremely unequal distribution of the combination of the carrier or non-carrier states for two SNPs (Fig. 6). Burfeind et al. (11) describe such a situation for rs3763040 and rs3763043. They observed that minor allele carriers of rs3763040 were most often also minor allele carriers of rs3763043, without the two SNPs being in linkage disequilibrium. A trend for one SNP might therefore be driven by the trend of the second SNP and the distribution of the carrier/non-carrier combinations of both. The discrepant findings for the effect direction (protective and deleterious) might therefore be a consequence of the presence of a second SNP with a rare carrier/non-carrier combination which is not adequately covered by the available data. Consequently, considering the joint effects of the AQP4 SNPs has the potential to yield more reliable associations.

**Figure 6.**
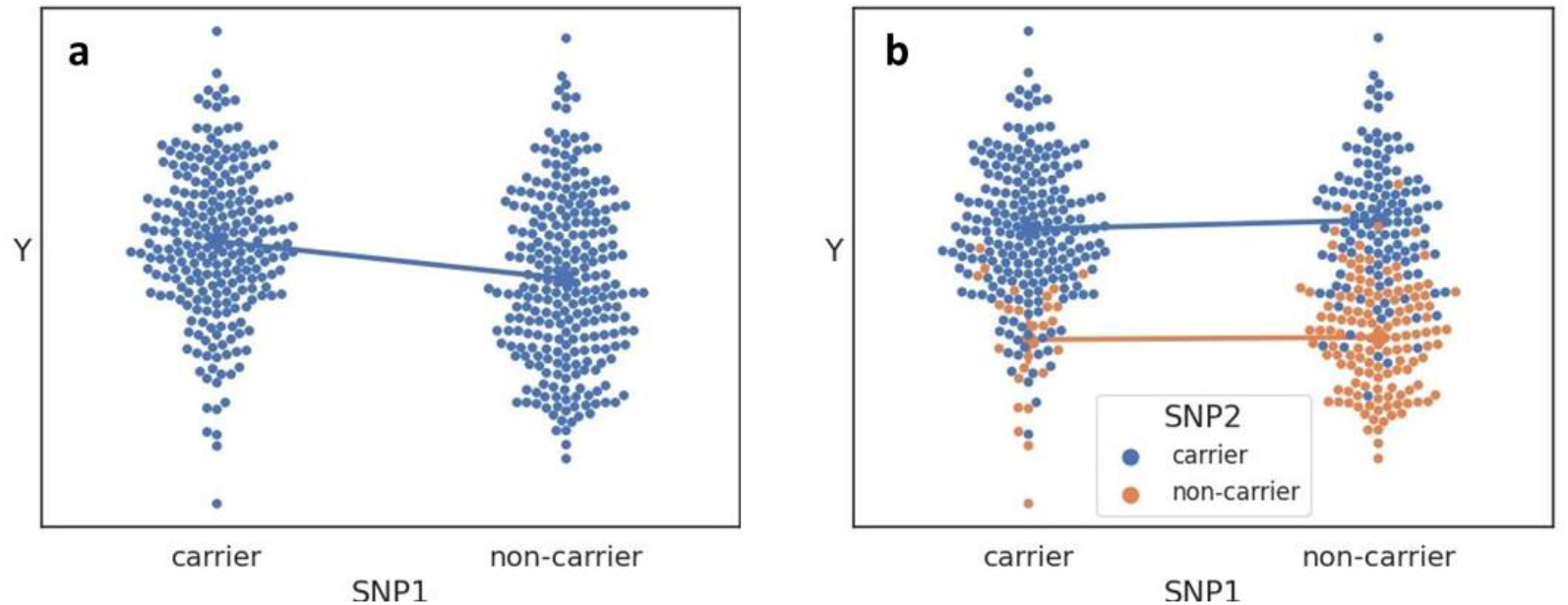
Simpson’s paradox: The trend for SNP1 in (a) disappears and even slightly inverses when the data are analysed in consideration of the status of SNP2 (b).This effect is a consequence of the extreme low amount of SNP2 non-carriers (orange dots) in SNP1 carriers. The two SNPs are not in linkage disequilibrium since SNP1 non-carriers can be both SNP2 carriers and non-carriers.

Besides effects for amyloid burden and disease progression in AD, SNPs of the risk score have also been found to be associated with sleep measures. rs335930 and rs16942851 are part of a haplotype which is associated with slow wave energy regulation in human NREM sleep (15). The SNP rs35248760 is in high LD with rs72878776, which has been found to be associated with poorer overall sleep quality, as well as to moderate the effect of sleep duration on brain amyloid burden (14). This provides a direct link between brain amyloid aggregation, sleep and AQP4. However, further studies are needed to understand the underlying molecular mechanisms.

While the studies of Chandra et al. and Burfeind et al. (11, 12) have found associations of genetic variations of *AQP4* with measures of dementia using classical linear models, these associations were mainly noted in cohorts of patients with significant clinical symptoms. With our method, which uses the joint effect of SNPs together with a stratification for sex, we were able to find associations which were consistent and reproducible in a cognitively unimpaired cohort. This may provide valuable insights into the disease’s underlying mechanisms at a very early stage in disease development, which opens up possibilities for early intervention or, potentially, prevention.

The joint effect of SNPs may be elucidated by considering their possible role as splicing enhancers or silencers in the transcription of AQP4 isoforms, providing a biological rationale for our method. Isoforms are protein variants that differ in the number of exons included during the transcription of genetic information. AQP4 can be translated by alternative splicing into the isoform AQP4-M1, consisting of exons 0 - 4 (see figure 1), which is freely mobile in the plasma membrane, or into the smaller isoform AQP4-M23, consisting of exons 1 – 4, which forms arrays in the astrocytic endfeet (13, 48). Recently a further isoform was described, an elongated variant named AQP4X, which is generated by a stop codon readthrough event and which is exclusively perivascular (49).

A limitation of our study is the small size of the dataset which was used for modelling and explanations, since rare combinations of SNP states could be missed. We used the smaller ADNI cohort instead of the much larger A4/LEARN cohort for modelling and explanations because, especially for the later visits, all diagnoses were present, from cognitively normal to AD, while in A4/LEARN only cognitively normal participants were included. A replication of our method with a larger dataset, also containing the whole bandwidth of disease, would be of high interest.

Our method also does not differentiate between epistatic and statistical interactions. Epistatic interactions describe a masking effect, whereby a status of a variant can be dominant over another variant in the sense that it prevents the other variant from manifesting its effect (19). Statistical interactions originate from the unequal distribution of the combinations of the carrier or non-carrier states of variants. The incorporation of a SNP-SNP-interaction specific model with consideration of epistatic interactions into the formulation of the risk score could have a potential to improve the performance of the risk score, and should be a topic of future investigations.

In summary, our results support the hypothesis of an involvement of the glymphatic system, and particularly AQP4, in the pathologic accumulation of ß-amyloid in the brain. Our findings suggest also that several *AQP4* SNPs show a synergistic and sex-dependent effect on early brain amyloid aggregation pathology, which can be expressed by an AQP4 polymorphism-based risk score. The risk score has the potential for practical application in stratification, such as in intervention studies or clinical trial recruitment. Additionally, it will be valuable for further investigations into associations with neuroinflammation and sleep, as well as into the role of AQP4 in amyloid aggregation.

## Data Availability

All data produced in the present study are available upon reasonable request to the authors.

https://ida.loni.usc.edu

## Acknowledgments

Data collection and sharing for this project was funded by the Alzheimer’s Disease Neuroimaging Initiative (ADNI) (National Institutes of Health Grant U01 AG024904) and DOD ADNI (Department of Defense award number W81XWH-12-2-0012), as well as by the A4 study (a4study.org).

ADNI is funded by the National Institute on Aging, the National Institute of Biomedical Imaging and Bioengineering, and through generous contributions from the following: AbbVie, Alzheimer’s Association; Alzheimer’s Drug Discovery Foundation; Araclon Biotech; BioClinica, Inc.; Biogen; Bristol-Myers Squibb Company; CereSpir, Inc.; Cogstate; Eisai Inc.; Elan Pharmaceuticals, Inc.; Eli Lilly and Company; EuroImmun; F. Hoffmann-La Roche Ltd and its affiliated company Genentech, Inc.; Fujirebio; GE Healthcare; IXICO Ltd.; Janssen Alzheimer Immunotherapy Research & Development, LLC.; Johnson & Johnson Pharmaceutical Research & Development LLC.; Lumosity; Lundbeck; Merck & Co., Inc.; Meso Scale Diagnostics, LLC.; NeuroRx Research; Neurotrack Technologies; Novartis Pharmaceuticals Corporation; Pfizer Inc.; Piramal Imaging; Servier; Takeda Pharmaceutical Company; and Transition Therapeutics. The Canadian Institutes of Health Research is providing funds to support ADNI clinical sites in Canada. Private sector contributions are facilitated by the Foundation for the National Institutes of Health (www.fnih.org). The grantee organization is the Northern California Institute for Research and Education, and the study is coordinated by the Alzheimer’s Therapeutic Research Institute at the University of Southern California. ADNI data are disseminated by the Laboratory for Neuro Imaging at the University of Southern California.

The A4 Study is funded by a public-private-philanthropic partnership, including funding from the National Institutes of Health-National Institute on Aging, Eli Lilly and Company, Alzheimer’s Association, Accelerating Medicines Partnership, GHR Foundation, an anonymous foundation and additional private donors, with in-kind support from Avid and Cogstate. The companion observational Longitudinal Evaluation of Amyloid Risk and Neurodegeneration (LEARN) Study is funded by the Alzheimer’s Association and GHR Foundation. The A4 and LEARN Studies are led by Dr. Reisa Sperling at Brigham and Women’s Hospital, Harvard Medical School and Dr. Paul Aisen at the Alzheimer’s Therapeutic Research Institute (ATRI), University of Southern California. The A4 and LEARN Studies are coordinated by ATRI at the University of Southern California, and the data are made available through the Laboratory for Neuro Imaging at the University of Southern California. The participants screening for the A4 Study provided permission to share their de-identified data in order to advance the quest to find a successful treatment for Alzheimer’s disease. We would like to acknowledge the dedication of all the participants, the site personnel, and all of the partnership team members who continue to make the A4 and LEARN Studies possible. The complete A4 Study Team list is available on: http://a4study.org/a4-study-team

